# Interleaved TMS/fMRI shows that threat decreases dlPFC-mediated top-down regulation of emotion processing

**DOI:** 10.1101/2023.11.11.23298414

**Authors:** Milan Patel, Marta Teferi, Abigail Casalvera, Kevin Lynch, Frederick Nitchie, Walid Makhoul, Desmond J Oathes, Yvette Sheline, Nicholas L Balderston

**Affiliations:** Center for Neuromodulation in Depression and Stress, Department of Psychiatry, University of Pennsylvania, Philadelphia, PA, USA; Center for Clinical Epidemiology and Biostatistics, Department of Psychiatry, University of Pennsylvania, Philadelphia, PA, USA

**Keywords:** dlPFC, interleaved TMS/fMRI, threat, anxiety, working memory, fMRI, TMS

## Abstract

**Background:** The right dorsolateral prefrontal cortex (dlPFC) has been indicated to be a key region in the cognitive regulation of emotion by many previous neuromodulation and neuroimaging studies. However, there is little direct causal evidence supporting this top-down regulation hypothesis. Furthermore, it is unclear whether contextual threat impacts this top-down regulation. By combining TMS/fMRI, this study aimed to uncover the impact of unpredictable threat on TMS-evoked BOLD response in dlPFC-regulated emotional networks. Based on the previous findings linking the dlPFC to the downregulation of emotional network activity, we hypothesized TMS pulses would deactivate activity in anxiety expression regions, and that threat would reduce this top-down regulation.

**Methods:** 44 healthy controls (no current or history of psychiatric disorders) were recruited to take part in a broader study. Subjects completed the neutral, predictable, and unpredictable (NPU) threat task while receiving TMS pulses to either the right dlPFC or a control region. dlPFC targeting was based on data from a separate targeting session, where subjects completed the Sternberg working memory (WM) task inside the MRI scanner.

**Results:** When compared to safe conditions, subjects reported significantly higher levels of anxiety under threat conditions. Additionally, TMS-evoked responses in the left insula (LI), right sensory/motor cortex (RSM), and a region encompassing the bilateral SMA regions (BSMA) differed significantly between safe and threat conditions. There was a significant TMS-evoked deactivation in safe periods that was significantly attenuated in threat periods across all 3 regions.

**Conclusions:** These findings suggest that threat decreases dlPFC-regulated emotional processing by attenuating the top-down control of emotion, like the left insula. Critically, these findings provide support for the use of right dlPFC stimulation as a potential intervention in anxiety disorders.

## Introduction

Difficulties in attention control, working memory, and cognitive processing are commonly experienced in individuals with anxiety disorders and these issues cut across diagnostic boundaries ^1–3^. The dorsolateral prefrontal cortex (dlPFC) is thought to be a key site in the brain’s cognitive control network, supporting cognitive processes like attention and working memory ^4–10^. Neuroimaging literature has also shown that this site is engaged during emotion regulation tasks, suggesting that emotion regulation is a key process mediated by the dlPFC ^11–18^. Accordingly, the dlPFC has become a common site for neuromodulatory intervention in both the clinic and the laboratory. However, the neuromodulation results often do not support theories of emotion regulation based on the neuroimaging findings, and further suggest that the degree to which the dlPFC supports emotion regulation may depend on the laterality of the activity ^19^.

While early neuroimaging findings supported the idea that the dlPFC contributed to emotion regulation, clinical data from repetitive transcranial magnetic stimulation (rTMS) began to question this assumption. TMS is a noninvasive form of neuromodulation that uses rapidly alternating electrical currents to generate magnetic fields that can activate brain cells. Depending upon the pattern and dose of stimulation, it is possible to either up- or down-regulate activity at the stimulation site. When applied clinically, patterns that up-regulate activity (high frequency rTMS or intermittent theta burst stimulation (iTBS)) tended to facilitate emotion regulation when applied to the left hemisphere ^19,20^. That is, they decreased symptoms of depression and anxiety. However, the results were different when the right dlPFC was stimulated. For stimulation applied here, it was the patterns of TMS that down-regulate activity that tended to facilitate emotion regulation ^19,20^. Specifically, 1 Hz stimulation to the right dlPFC has been shown to reduce anxiety in individuals with comorbid depression and anxiety. However, the mechanism is unclear.

One way to understand how right dlPFC activity contributes to emotion regulation is to stimulate the right dlPFC and measure the effect of this stimulation on activity in downstream regions using interleaved TMS/fMRI. During interleaved TMS/fMRI studies, single pulses are administered to a stimulation site during pre-programmed gaps in the fMRI sequence, allowing the experimenter to measure TMS-evoked BOLD activity in regions that are downstream of the stimulation site ^21,22^. Such an approach allows researchers to causally map the networks associated with the stimulation site.

In this study we used interleaved TMS/fMRI to investigate the downstream responses following right dlPFC stimulation during periods of elevated anxiety. Subjects completed the neural, predictable, unpredictable (NPU) threat task while undergoing TMS/fMRI ^23–25^. During this task, subjects were instructed to rate their anxiety during periods of predictable and unpredictable threat. Periodic pulses were delivered to either the right dlPFC or a control region during the different blocks. According to the regulation hypothesis, we should expect to see downregulation threat processing, resulting in decreases in of BOLD activity in threat-related regions when stimulating the right dlPFC during threat. In contrast, according to the expression hypothesis we should expect to see up-regulation of threat processing, resulting in increases in of BOLD activity in threat-related regions when stimulating the right dlPFC during threat.

## Materials and Methods

### Participants

Sixty-eight right-handed participants between the ages of 18 and 50 were recruited from Philadelphia, PA, to take part in the broader study. A total of 44 participants elected to complete the optional TMS/fMRI visit described below. A total of 41 participants completed all aspects of the study needed for the current work (31 females, 13 males, mean age□=□ 25.39 years, SD□=□6.55). Exclusion criteria included: current or past Axis I psychiatric disorder(s) as identified with the Structured Clinical Interview (SCID) for DSM-V (Research Version) ^26^, use of psychoactive medications, any significant medical or neurological problems (e.g. cardiovascular illness, respiratory illness, neurological illness, seizure, etc.), and any MRI/TMS contraindications (e.g. implanted metal, history of epilepsy or seizure, etc.). For a complete list, see: www.clinicaltrial.gov (Identifier: NCT03993509). Three subjects were excluded due to technical issues. All participants signed an informed consent form, and the protocol was approved by the Institutional Review Board for human subject research at the University of Pennsylvania. All procedures contributing to this work comply with the ethical standards of the relevant national and institutional committees on human experimentation and with the Helsinki Declaration of 1975, as revised in 2008.

### General Procedure

#### General

Participants were enrolled in a broader study where they received multiple sessions of active or sham cTBS or iTBS, followed by a series of cognitive and behavioral tests. The results of this broader study are described elsewhere. The current results come from an optional TMS/fMRI session included in this broader study where subjects completed the neutral predictable and unpredictable (NPU) threat task while receiving periodic TMS pulses to either the right dlPFC or a control region.

On the consent visit, subjects completed the consent form, the MRI safety form, the TMS adult safety screen (TASS), and a medical history questionnaire, and the coordinator administers the SCID, Montgomery-Asberg Depression Rating Scale (MADRS) ^27^, and an eligibility checklist. Subjects who meet screening criteria continued to complete the demographics questionnaire, the State/Trait Anxiety Inventory (STAI) ^28^, and the Beck Anxiety Inventory (BAI) ^29^. During the MRI targeting visit, subjects were escorted to the scanner and given ear plugs, a button box, an emergency squeeze ball, and padding to minimize head movement. Next, anatomical scanning was completed from start to finish. Finally, subjects then completed 1 run of the Sternberg WM task and 2 resting state runs. During the first TMS visit of the broader study, motor threshold was established.

During the TMS/fMRI sessions, subjects were given a swimcap and the shock electrodes. Next the subjects head was registered with their MRI in Brainsight and their stimulation sites were marked on the swimcap. Afterward, the subjects completed the shock workup procedure and were escorted to the scanner. Once prepped for the first TMS/fMRI scan, the coil was positioned over the right dlPFC stimulation site and the articulating arm was supported with additional padding. Next the subjects completed 2 runs of the NPU task with TMS pulses targeting the right dlPFC. Afterward, the subjects were removed from the scanner and the coil was repositioned over the control site, such that their head was resting on the TMS coil and secured with additional padding. The subject then completed 2 additional NPU runs with the TMS pulses targeting the IPS.

### Materials

#### NPU task

Subjects completed two runs of the NPU task for each stimulation site during the TMS/fMRI session. Each run consisted of alternating blocks of neutral, predictable, and unpredictable blocks. During the neutral blocks, subjects were at no risk of being shocked. During the predictable blocks, subjects were only at risk of shock during the visual cue. During the unpredictable blocks, subjects were at risk of shock throughout. Threat blocks were always separated by a neutral block, resulting in the following two block orders: NPNUNUNP or NUNPNPNU. Like the laboratory version of the NPU task, which uses white noises to probe the acoustic startle reflex, our version consisted of “cue” and “intertrial interval (ITI)” trials where a TMS burst was administered during the presence or absence of a visual cue. These cues were simple colored shapes that varied across conditions. Each neutral block included 2 trials per condition, while each threat block included 4 trials per condition, totaling to 8 trials per condition per run. Three shocks were presented on each run at random points during either the cue (predictable condition) or the ITI (unpredictable condition) trials. Throughout the task subjects rated their anxiety from a scale of 0 (feeling not anxious) to 10 (feeling extremely anxious) using an onscreen numerical scale.

#### Sternberg WM targeting task

Subjects were presented with a series of maintain and sort trials. Each trial started with an instruction prompt to indicate the trial type, followed by a series of 5 letters, presented sequentially. These letters were retained in working memory for a brief interval. Afterward, subjects were given a forced choice response. On “maintain” trials, subjects were instructed to remember the letters in the order presented. On “sort” trials, subjects were instructed to rearrange the letters in alphabetical order. At the forced choice prompt, subjects viewed a letter/number combination, and were instructed to indicate whether the position of the letter in the series matched the number. Half of the trials were matches and the other half were mismatches. The duration of the letter series (1.5 – 2.5□s), retention interval (6.5 – 8.5□s), and ITI (5 – 8□s) were jittered across trials. The duration of the instructions (1 s) and response prompt (3 s) were fixed. There were 12 trials each for the sort and maintain conditions.

#### Shock

The shock stimulus consisted of a 100 ms train of 2 ms pulses delivered at 200 Hz using a using a DS7A constant current stimulator (Digitimer #DS7A, Ft. Lauderdale, FL). Shocks were delivered to the subject’s left wrist via disposable 11 mm Ag/AgCl electrodes (Biopac Item number EL508; Goleta, CA), spaced ∼2 cm apart. The shock intensity was calibrated prior to the TMS/fMRI session using an individualized workup procedure where subjects rated a series of shocks shock on a scale from 1 (not uncomfortable) to 10 (uncomfortable but tolerable) until the subject reached their “level 10”. Shocks during the session were delivered at the level.

#### Scans

MRI data were acquired using a 3 Tesla Siemens Prisma scanner. Structural scans used a 64-channel head coil (Erlangen, Germany). Structural scans included a T1-weighted MPRAGE (TR□=□2200□ms; TE□=□4.67□ms; flip angle□=□8°) with 160, 1□mm axial slices (matrix□=□256□ ×□256; field of view (FOV)□=□240□mm□×□240□mm), and a T2-weighted image (TR□=□3200□ms; TE□=□563□ms; flip angle□=□variable) with 160, 1□mm sagittal slices (matrix□=□256□mm□×□256□mm; FOV□=□240□mm□×□240□mm). TMS/fMRI scans used a single-channel birdcage coil (RAPID quad T/R single channel; Rimpar, Germany). Each run included 233 whole-brain BOLD images (TR□=□2000 ms; TE□=□30□ms; flip angle□=□75°) comprised of 32, 4□mm axial slices (matrix□=□64□×□64; FOV□=□192□mm□×□192□mm) aligned to the AC-PC line.

#### TMS/fMRI Pre-processing

TMS/fMRI data were processed using the afni_proc.py script distributed with the AFNI software package ^30^. The following the following preprocessing blocks: `tshift, align, tlrc, volreg, blur, mask, scale, regress` were used. During preprocessing, 1) the images were slice time corrected, 2) aligned to the T1 data using an Local Pearson Correlation cost function, 3) normalized to the MNI152_2009 templated distributed with AFNI, 4) individual volumes were registered to the image with the fewest outliers, 4) images were resampled to 3 mm isotropic voxels and blurred with a 6 mm Gaussian kernel, 5) masked using the union of the EPI brain mask and the skull-stripped T1, 6) scaled so the mean of each voxel timeseries was 100. The first 4 TRs and TRs with greater than 0.5 mm displacement or greater than 30% of voxels registered as outliers were censored/scrubbed from the timeseries prior to the GLM. The subject-level GLM included a set of polynomial regressors to model the baseline and regressors of no interest corresponding to the 6 primary motion vectors and their derivatives. NPU events were modeled with variable duration blocks to account for jittering in the timing of the events. TMS bursts were modeled using an impulse response function.

#### Target localization

Data from the Sternberg WM task was used to identify the right dlPFC target coordinates for the TMS/fMRI session ^24,31^. BOLD maps from the retention interval were masked with a function ROI of the right dlPFC. The ROI was obtained from a group-level analysis using a previously collected Sternberg WM dataset ^32,33^. Single subject BOLD activity was contrasted across sort and maintain trials and the coordinates for the peak voxel within this mask was extracted and used as a target. For the control site, we used the group level coordinates for the right intraparietal sulcus, identified in a previous study ^34,35^. We used the Brainsight (Rogue Research Inc, Montreal, Canada) frameless stereotaxic neuronavigation system to mark the target site on a swim cap worn during the TMS/fMRI session.

#### Motor threshold determination

A Magventure MagPro 100X stimulator with a B65 figure-8 coil was used to obtain resting motor threshold. Motor threshold was determined from EMG recordings taken from the first dorsal interosseous muscle (FDI) using the adaptive parameter estimation by sequential testing (PEST) algorithm ^36^. Stimulator output during the TMS/fMRI sessions was adjusted to account for differences in coil output.

#### Active stimulation

A Magventure MagPro 100X stimulator with a B91 figure-8 coil was used for the TMS/fMRI session. Periodically during the NPU task, subjects received single 3-pulse 50 Hz bursts at 100 percent of motor threshold adjusted for differences in coil output.

### Data analysis

#### Anxiety ratings

Anxiety ratings at the onset of each TMS burst were extracted and averaged across all trials in the safe and threat conditions. These responses were then compared using a paired sample t-test.

#### Whole brain TMS evoked responses

The first-level beta coefficients were extracted for all dlPFC-targeted TMS bursts. To identify TMS-evoked responses, we compared these betas to an implicit baseline using a single-sample t-test against 0 (`3dttest++`). To correct for multiple comparisons we used a cluster-based thresholding approach implemented by `3dClustSim` ^37^. We ran 10,000 Monte Carlo simulations with a voxelwise p-value of 0.001, a non-Gaussian (i.e. autocorrelation function) ^38^ estimation of the smoothness of the BOLD data, and extracted clusters comprised of voxels with adjoining faces or edges. Based on these simulations, we selected a minimum cluster size of 40, 3-mm voxels, which corresponded to a cluster-level p-value < 0.01.

#### Effect of threat on TMS-evoked BOLD responses

We extracted the average dlPFC-targeted TMS-evoked BOLD response during the safe and threat conditions for each of the clusters identified in the whole brain analysis. We then compared these responses using a paired sample t-test. We repeated this process for our control site evoked responses in a subset of individuals who also had data targeting the right IPS.

## Results

### Ratings

As a manipulation check, we compared the anxiety ratings in safe compared to threat blocks using a t-test. As expected, subjects reported significantly greater anxiety during threat periods compared to safe periods (t(39) = 8.85; p < 0.001; d = 1.4).

### TMS-evoked responses

We began by defining a set of regions that were directly activated by the right dlPFC TMS pulses by collapsing across conditions and computing a voxelwise t-test against zero. We used cluster thresholding and *monte carlo* simulations to correct for multiple comparisons. We identified 7 regions with TMS-evoked responses that significantly differed from zero (Figure 3 A-G, Table 1). To determine whether the TMS-evoked responses in these regions differed as a function of threat, we split trials into safe and threat conditions and performed paired sample t-tests. We found that TMS-evoked responses in the right sensory/motor cortex (Figure 3 H “RSM”), the left insula (Figure 3 H “LI”), and a region encompassing the bilateral SMA regions (Figure 3 H “BSMA”) differed significantly in the threat compared to safe conditions. Across all 3 regions, there was a significant TMS-evoked deactivation in the safe periods that was significantly attenuated in the threat periods (RSM: t(40) = 2.29; p = 0.027; d = 0.36; LI: t(40) = 2.27; p = 0.029; d = 0.37; BSMA: t(40) = 2.25; p = 0.03; d = 0.36). No other region showed a significant differentiation as a function of threat (all p-values > 0.05).

**Figure 1.**
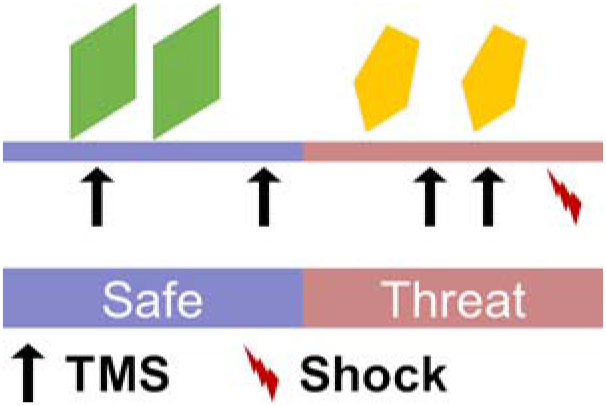
Schematic of task design. We extracted data from safe and (unpredictable) threat periods during the NPU paradigm. During the safe period, subjects could not receive a shock (red lightning). During the threat periods, subjects were instructed that they could receive a shock at any time. TMS pulses (arrows) were presented at random intervals throughout the task during both cue (shapes) and ITI periods.

**Figure 2.**
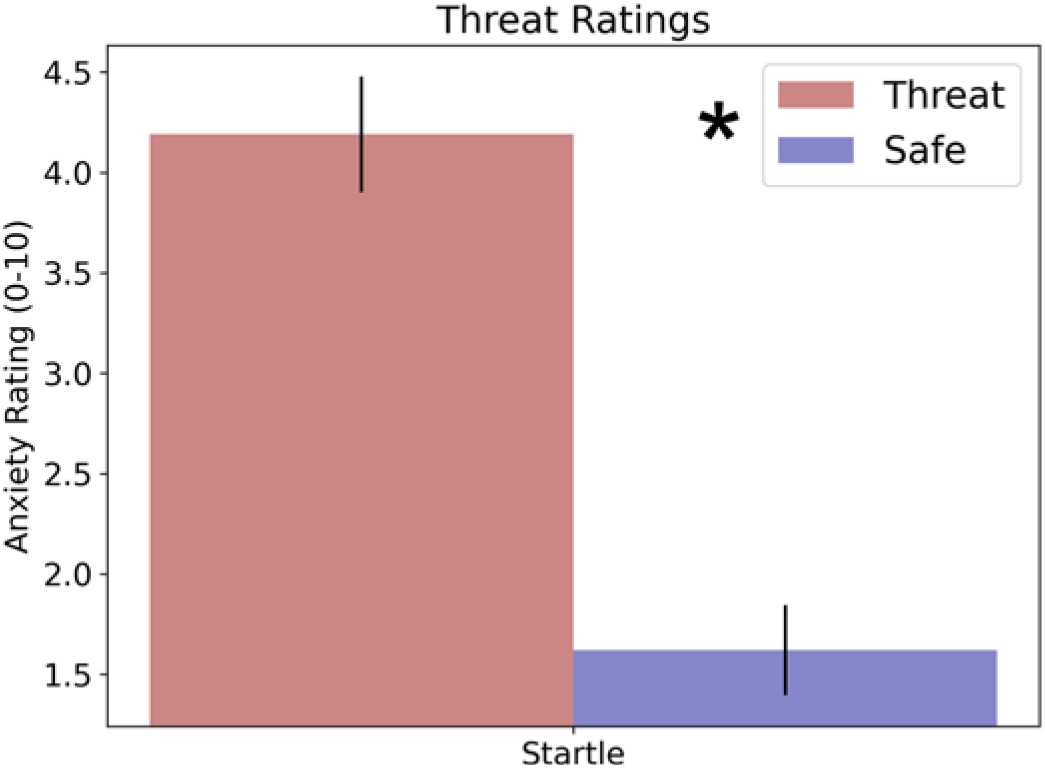
Anxiety ratings threat task. Anxiety ratings reported on a scale from 0 to 10. Bars represent the mean ± SEM. * = p < 0.05.

**Figure 3.**
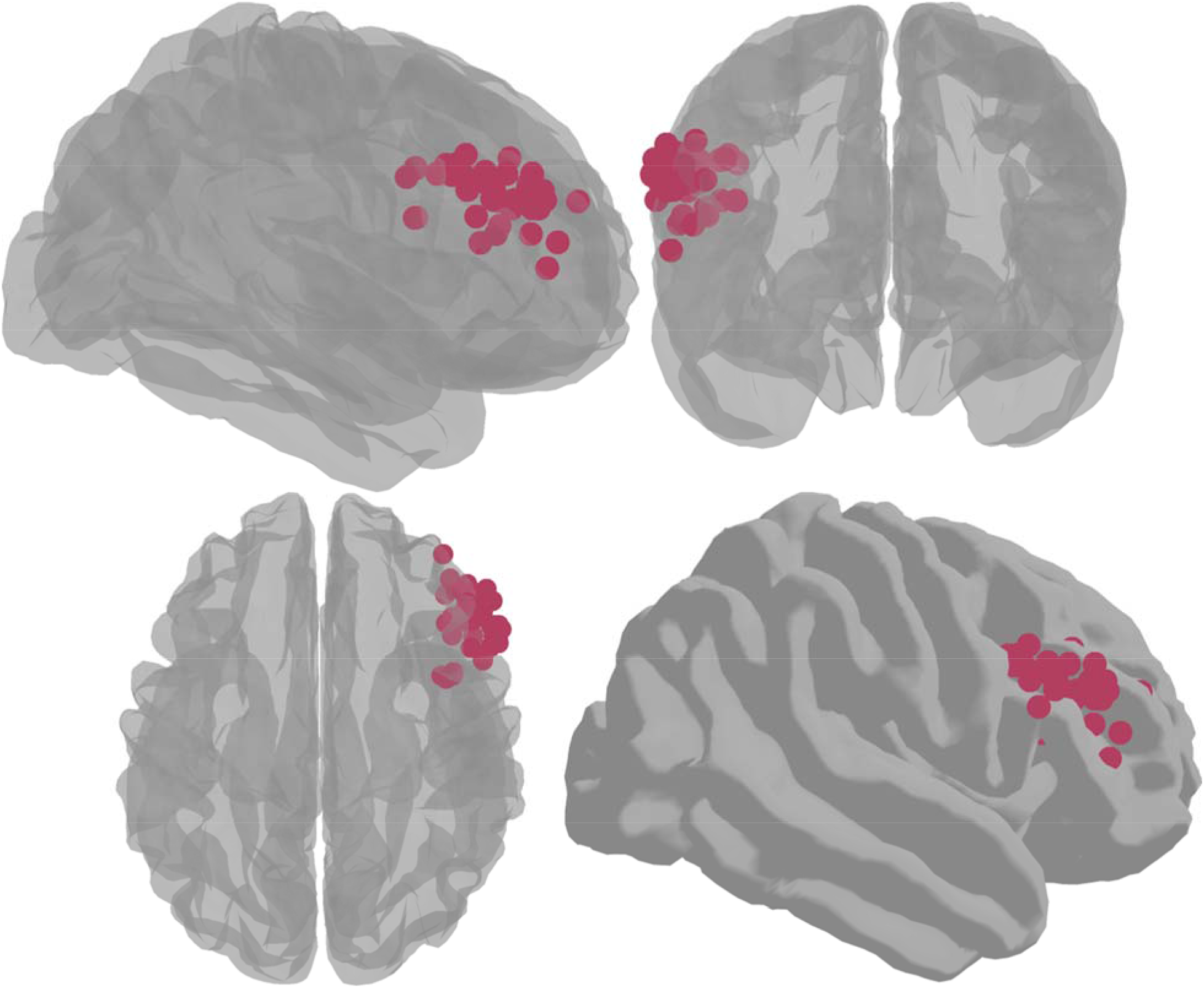
Single subject targets in MNI space. Spheres represent the single-subject peaks for WM-related activity during the Sternberg WM task, which were used as the TMS targets during the TMS/fMRI scans.

**Figure 4.**
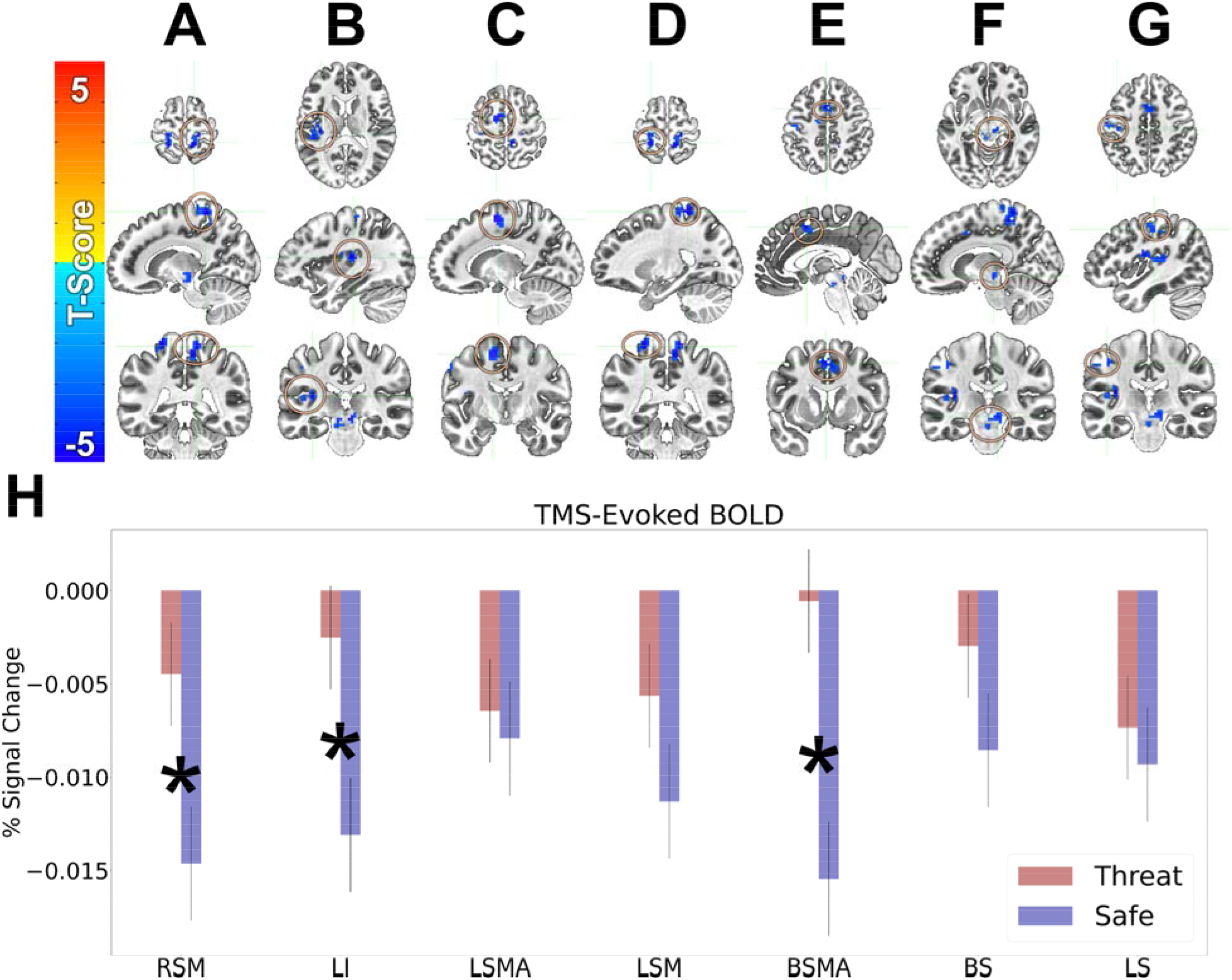
BOLD responses evoked by TMS pulses delivered to the right dlPFC. A-G) Regions showing TMS-evoked BOLD responses that significantly differ from an implicit baseline. H) BOLD responses in these regions plotted as a function of safe and threat conditions. RSM = Right Sensory/Motor. LI = Left Insula. LSMA = Left SMA. LSM = Left Sensory/Motor. BSMA = Bilateral SMA. BS = Brainstem. LS = Left Sensory. Warm colors represent activations. Cool colors represent deactivations. Bars represent the mean ± SEM. * = p < 0.05.

**Figure 5.**
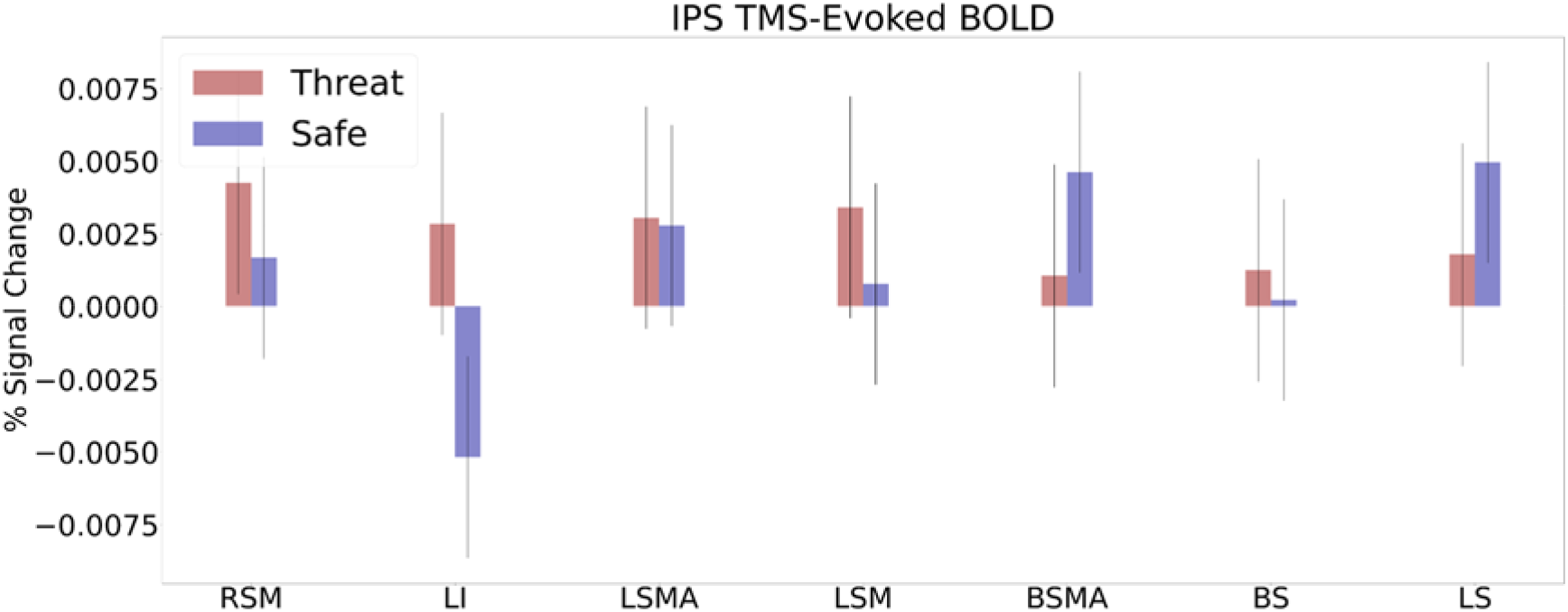
BOLD responses evoked by TMS pulses delivered to a control site. BOLD responses in the regions from Figure 4 evoked by TMS pulses delivered to a control site, plotted as a function of safe and threat conditions. RSM = Right Sensory/Motor. LI = Left Insula. LSMA = Left SMA. LSM = Left Sensory/Motor. BSMA = Bilateral SMA. BS = Brainstem. LS = Left Sensory. Warm colors represent activations. Cool colors represent deactivations. Bars represent the mean ± SEM.

### Control site-evoked responses

To determine whether the differential responses evoked by the dlPFC targeted TMS pulses were specific to dlPFC stimulation, we examined the responses in these same regions following stimulation to a control site (the right IPS) in a subset of participants. None of these regions showed significant differentiation as a function of threat. Indeed, the site with the largest effect size showed a Cohen’s d of 0.22, which would require ∼130 subjects to reliably detect an effect.

## Discussion

In this study, we investigated the effect of threat on TMS-evoked BOLD responses targeted at the right dlPFC. By administering TMS pulses interleaved with the fMRI acquisition, we were able to measure BOLD responses downstream from the dlPFC stimulation site. By nesting these TMS pulses in alternating periods of safety and threat, we were able to determine how threat effected activity in regions downstream from the dlPFC. During safe periods, we found that right dlPFC stimulation significantly deactivated a variety of regions across several brain networks. However, during the threat periods, this deactivation was attenuated in regions like the anterior insula that are specifically involved in threat processing. Together these results suggest that the dlPFC plays a broad role in the top-down suppression of network activity, which could help to filter distracting information out of working memory. However, during periods of elevated threat and arousal, this top-down suppression is reduced, allowing for an increased vigilance to possible threats in the environment.

The dlPFC is known to play a key role in working memory processes and emotion regulation ^19^. However, the link between these two aspects of dlPFC function is currently unclear. Recently, we proposed a model whereby the primary role of the dlPFC is to subserve working memory related functions like the maintenance, manipulation, and suppression of items in short term stores ^23,24,31,39^. Furthermore, we proposed that the left and right dlPFC were specialized to process distinct domains of content in this processing ^19,40^. According to this model, the left dlPFC is specialized for verbal content, while the right dlPFC is specialized for non-verbal content. Accordingly, the left and right dlPFC could potentially play distinct roles in emotion regulation, with the left mediating effortful verbal regulation strategies, and the right mediating more automatic non-verbal strategies. The current results fit within this model and suggest that external stimulation of the right dlPFC may trigger this automatic emotion regulation by suppressing activity in emotion related regions during safe periods and to a lesser extent during threat periods. It could be argued that this attenuation of emotion regulation during threat is adaptive, allowing for the expression of context appropriate fear responses.

Clinical anxiety disorders encompass a constellation of symptoms including hyperarousal, hypervigilance, impaired attention control, and overgeneralization ^41,42^. Many of these symptoms could be explained using this working memory framework. For instance, hypervigilance and impaired attention control could arise from impaired top-down inhibition of distraction-related activity when cognitive demands are high ^2,41^. Similarly, overgeneralization of threat could occur when novel memory encoding events occur during periods of elevated arousal that are mediated by this impaired top-down inhibition ^43,44^. These memories, in non-anxious individuals would typically be recorded as neutral events, but instead they could acquire a negative valence in anxious individuals.

The dlPFC is currently one of the most widely used targeting sites for rTMS in depression patients, and there is good evidence to suggest that left dlPFC stimulation potentially improves depression symptoms by normalizing connectivity between the dlPFC and the subgenual anterior cingulate cortex ^20,39,45^. However, the mechanisms that mediate anxiety reduction following right dlPFC stimulation. Indeed, it is even unclear what type of stimulation should offer the best results. There is some evidence that 1 Hz stimulation to the right dlPFC can reduce anxiety symptoms in depressed individuals. There is also some evidence that 5 Hz and iTBS to the right dlPFC can improve symptoms of PTSD. While there is no clear cut relationship between stimulation type and excitability, it is generally thought that 1 Hz is potentially inhibitory, while 5 Hz and iTBS are potentially excitatory, making it difficult to suggest that these clinical stimulation protocols are targeting similar mechanisms. Similarly, as part of this larger project, we have administered several sessions of either cTBS or iTBS and measure the effect of these stimulation sessions on anxiety potentiated startle. Contrary to our hypotheses, both cTBS and iTBS increased anxiety potentiated startle, further complicating our attempts at deriving a comprehensive mechanistic explanation of the link between dlPFC stimulation and anxiety. It should be noted that our studies were conducted in low anxious healthy individuals and may not generalize to patient populations. Still, one might expect distinct effects for the differing theta burst protocols. Future research is needed. Suffice to say that our current finding that external stimulation of the right dlPFC directly inhibits activity in downstream regions will be important for disentangling these effects.

### Strengths and Limitations

By combining TMS and fMRI techniques, this study was able to simultaneously stimulate the right dlPFC and record brain activity in related regions, providing direct evidence that right dlPFC activity can downregulate activity in downstream regions. Despite this innovation, the study has several limitations. First, by using an active control region, it is difficult to disentangle the effects of the target and the control site. We accounted for this by analyzing the control site data separately and showing that the pattern of results differed from those evoked by active stimulation of our dlPFC target. However, future work should incorporate the use of a realistic sham condition. Additionally, of the 68 initially recruited participants, only 44 completed the TMS/fMRI sessions, and because the control condition was added later, we do not have data from all subjects collected from this region. Future work should extend these results and test their implications in a larger sample. Finally, although we induced anxiety using threat of unpredictable shock, this paradigm is not a stand-in for clinical anxiety. Future work should include anxious subjects in the sample.

### Conclusions

In summary, the results of this study supported our original hypothesis that threat would impact dlPFC-regulated anxiety expression. The introduction of threat conditions resulted in decreased activity in downstream regions, such as the left insula, which indicates the dlPFC regulates anxiety expression through downregulating downstream activity. These results not only suggest a relationship between the dlPFC and left insula in anxiety regulation, but also suggests right dlPFC stimulation can be a reliable targeting spot for anxiety regulation. Further research can expand these implications to better understand the neural mechanisms underlying anxiety regulation during threat.

## Data Availability

All data produced in the present study are available upon reasonable request to the authors

## Author contributions

CRediT author statement according to: https://www.elsevier.com/authors/policies-and-guidelines/credit-author-statement.

**Conceptualization**: DJO, YIS, NLB

**Methodology**: DJO, NLB

**Software**: NLB

**Formal analysis**: KL, NLB

**Investigation**: MP, MT, AC, FN, WM, NLB

**Writing - Original Draft**: MP, NLB

**Writing - Review & Editing**: MP, MT, AC, FN, WM, KL, DJO, YIS, NLB

**Visualization**: MP, KL, NLB

**Supervision**: NLB

**Project administration**: NLB

**Funding acquisition**: NLB

## Acknowledgments

The study team would like to thank the following individuals who contributed to Dr. Balderston’s K01 project: Dr. Kerry Ressler, Dr. Michael Thase, and Dr. Kristin Linn. The study team would like to also thank the DSMB members who oversaw the project: Dr. Lindsay Oberman (Chair), Dr. Alex Shackman, Dr. Gang Chen. This study utilized the high-performance computational capabilities of the CUBIC computing cluster at the University of Pennsylvania. (https://www.med.upenn.edu/cbica/cubic.html). The authors would like to thank Maria Prociuk for her expertise and assistance in submitting the paper. We would also like to thank the participants for their time and effort.

## Disclosures

This project was supported in part by 2 NARSAD Young Investigator Grants from the Brain & Behavior Research Foundation (NLB: 2018, 2021); and by a K01 award K01MH121777 (NLB). The authors report no biomedical financial interests or potential conflicts of interest.

## Ethical Standards

The authors assert that all procedures contributing to this work comply with the ethical standards of the relevant national and institutional committees on human experimentation and with the Helsinki Declaration of 1975, as revised in 2008.

## Notes

### Competing Interest Statement

The authors have declared no competing interest.

### Clinical Trial

NCT03993509

### Clinical Protocols

https://clinicaltrials.gov/ct2/show/record/NCT03993509

### Author Declarations

All participants signed an informed consent form, and the protocol was approved by the Institutional Review Board for human subject research at the University of Pennsylvania. All procedures contributing to this work comply with the ethical standards of the relevant national and institutional committees on human experimentation and with the Helsinki Declaration of 1975, as revised in 2008.

